# Exome Reanalysis Identifies Novel Candidate Genes Associated with Congenital Anomalies of the Kidney and Urinary Tract in China

**DOI:** 10.64898/2026.02.03.26345078

**Authors:** Huiru Sun, Chunyan Wang, Wentao Zhang, Mingji Deng, Qian Shen, Jianhua Mao, Qing Sun, Huimin Luo, Huijun Shen, Jingjing Wang, Dandan Xin, Yan Zhou, Miaomiao Li, Yihui Zhai, Ying Cao, Hong Xu, Shaohua Fan, the Chinese Children Genetic Kidney Disease Database (CCGKDD) investigators

**Affiliations:** Department of Nephrology, State Key Laboratory of Genetics and Development of Complex Phenotypes, Center for Evolutionary Biology, Children’s Hospital of Fudan University, School of Life Sciences, Human Phenome Institute, Fudan University, Shanghai, 200438, China; State Key Laboratory of Cardiology and Medical Innovation Center, Shanghai East Hospital, School of Life Sciences and Technology, Tongji University, Shanghai, 200092, China; Department of Nephrology, The Children’s Hospital of Zhejiang University School of Medicine, Hangzhou, 310003, China; Department of Nephrology and Rheumatology, Women and Children’s Hospital, Qingdao University, Qingdao, 266035, China; Department of Nephrology, The First People’s Hospital of Yunnan Province, Kunming, 650032, China

**Keywords:** congenital anomalies of the kidney and urinary tract, pediatric genetic kidney disease, exome sequencing, exome reanalysis, multicenter database, F0 knockout zebrafish, renal development

## Abstract

Congenital anomalies of the kidney and urinary tract (CAKUT) are the primary cause of pediatric kidney failure, yet the genetic etiologies remain elusive for most affected individuals. Reanalysis of trio exome sequencing data from 80 Chinese CAKUT patients identified 32 rare, predicted deleterious variants. Replication in unrelated families from a national multicenter database prioritized four novel candidate genes—*DOCK11*, *MIB1*, *TENM2*, and *TNS1*. These candidates are involved in both well-characterized developmental pathways and more under-explored biological processes relevant to renal and ureteric morphogenesis. CRISPR-Cas9-mediated zebrafish knockout studies were employed to validate the potential association of these genes with kidney abnormalities including significant pericardial edema, malformed renal tubules, and impaired glomerular filtration. These findings offer potential genetic diagnoses for 10% of CAKUT probands, and demonstrate that exome reanalysis can substantially improve diagnostic yield and inform personalized clinical management. Overall, this study expands the known genetic landscape of CAKUT.

## Introduction

Congenital anomalies of the kidney and urinary tract (CAKUT) represent a heterogeneous group of developmental disorders affecting the kidney and/or urinary tract, and are a leading cause of kidney failure in children and adolescents(*1–4*). The clinical spectrum of CAKUT is broad, encompassing abnormalities in number (e.g., renal agenesis, duplex kidney), size (e.g., renal hypoplasia), shape (e.g., horseshoe kidney), position (e.g., ectopic kidney and ureter), tissue structure (e.g., vesicoureteral reflux (VUR)), and tissue composition (e.g., renal dysplasia) of the kidney and/or urinary tract(*4*, *5*). Secondary upper urinary tract damage, such as hydronephrosis, may arise from more distal lesions, including ureterovesical junction obstruction or VUR(*4*, *6*). While most cases of CAKUT are ‘isolated’, 6–50% of patients occasionally present with ‘syndromic’ CAKUT, in which renal anomalies co-occur with extra-renal malformations, such as cardiac, vertebral, limb or ocular defects(*7–9*). CAKUT manifestations fall within the phenotypic spectrum of over 200 syndromes(*3*, *10*), contributing to considerable phenotypic variability.

Globally, CAKUT is the most common cause of end-stage kidney disease (ESKD) in children and adolescents, accounting for around 20–30% of all congenital anomalies(*1*, *11–14*). It occurs with an estimated frequency of 4–60 per 10,000 live births(*15–17*). In China, 33,621 hospital patients were diagnosed with CAKUT between January 1, 2016, and December 31, 2022, with a male-to-female ratio of approximately 1.9:1(*5*). Hydronephrosis was the most common presentation (27.84%), and the proportion of CAKUT cases among total hospitalizations increased steadily during this period, indicating a rising disease burden(*5*). In addition, the prevalence of syndromic CAKUT, such as in cases of 17q12 microdeletion, may be underestimated(*4*, *18*).

A range of pathogenic or likely pathogenic variants has been implicated in the pathogenesis of CAKUT, affecting genes encoding proteins involved in critical developmental pathways. These include transcription factors (e.g., *PAX2*, *HNF1B*), signaling molecules (e.g., *ROBO*2), and extracellular matrix components of the developing kidney (e.g., *FRAS1*, *FREM2*, *ITGA8*)(*19*, *20*). Several signaling pathways implicated in CAKUT pathogenesis, particularly in ureteric bud (UB) branching, include glial-derived neurotrophic factor (GDNF)–RET(*21*, *22*), retinoic acid (*1*, *23–25*), Notch(*26*, *27*), phosphoinositide 3-kinase-AKT (PI3K-Akt)(*28*, *29*), mitogen-activated protein kinase (MAPK)(*30*, *31*), as well as cyclic adenosine 3′,5′-monophosphate (cAMP)(*32*, *33*) signaling pathways. While some genetic variants are linked to isolated CAKUT phenotypes, other alleles in the same gene cause syndromic phenotypes involving the skeletal system(*34*), eyes(*35*), and/or brain(*36–38*). Such pleiotropy contributes to the variable expressivity and complex genotype-phenotype correlations observed in familial studies(*4*, *39*, *40*), making the identification of novel CAKUT genes and proving causality particularly challenging(*4*, *41*).

Recent advances in next-generation sequencing technologies, including targeted gene panels and exome sequencing (ES), have revolutionized the studies on CAKUT etiologies(*42*). To date, 54 monogenic causes have been implicated in isolated or syndromic CAKUT(*4*). In addition, 131 gene candidates have been associated with syndromes in which CAKUT features appear as facultative manifestations (occurring in fewer than half of the reported cases)(*4*) (**Supplementary Table 1**). However, genetic testing has achieved diagnostic rates of just 6-20% in pediatric cohorts(*43–52*), with the majority of these studies relying on samples of European ancestry(*4*, *53*). The persistent diagnostic gap, underrepresentation of diverse populations, and intricate genotype-phenotype associations highlight an urgent need to identify novel candidate genes and refine molecular diagnostic strategies for CAKUT.

This study analyzed ES data from 80 probands and their parents, representing a spectrum of CAKUT phenotypes—including renal agenesis, renal hypo/dysplasia (RHD), renal cystic dysplasia, hydronephrosis, VUR, ectopic or horseshoe kidney, duplex kidney, and posterior urethral valves—recruited from multiple medical centers across China. All participants had previously received negative results from routine clinical genetic testing. Trio-based ES was employed to identify potential disease-causing variants and candidate genes in probands, followed by functional validation in F0 zebrafish. This integrated approach enabled the elucidation of candidate genetic contributors to CAKUT in the Chinese population, potentially improving diagnostic yield and informing personalized management.

## Results

### Demographics and clinical data

In this study, we analyzed 80 probands with suspected CAKUT who had previously received negative diagnostic evaluations. These cases were sourced from the Chinese Children Genetic Kidney Disease Database (CCGKDD)(*49*), a national network (cohort sample size: 4,140) dedicated to cataloging phenotypic and genetic information of patients suffering from five major kinds of kidney disease, including CAKUT, glomerulopathy, renal cystic disease, and renal tubular disorders, among others.

The 80 samples in the final cohort had an age at diagnosis ranging from prenatal stages to 14.4 years, with a median age of 3.0 (IQR: 0.3–8.0) years. Of these, 58 patients (72.5%) were male and 22 (27.5%) were female. 42.5% (n = 34) of the CAKUT occurrences were observed during the newborn period (n = 17, birth to 2 months) and the school-age period (n = 17, 8–12 years) (**Supplementary Table 2**).

Among the 80 diagnosed cases, a wide spectrum of renal and ureteral malformations was observed, including renal parenchymal defects (RHD (n = 45), renal cystic dysplasia (n = 12), and agenesis (n = 3)), anomalies of the collecting system (hydronephrosis (n = 12) and duplex kidney (n = 1)), and positional or fusion abnormalities (renal ectopia or horseshoe kidney (n = 4)). RHD was the most represented phenotype in association with VUR and/or other urinary abnormalities (such as hyperuricemia and posterior urethral valve). Isolated VUR were observed in 10 cases. Notably, 76.3% of cases (n = 61) exhibited bilateral involvement. In addition, dual renal and ureteral phenotypes were observed in seven probands, six of whom presented with hydronephrosis, which may reflect secondary effects of renal malformations or concomitant urinary tract involvement. Overall, 56.3% (45/80) presented with isolated kidney abnormalities, whereas 43.7% exhibited syndromic manifestations, including deafness, seizures, scoliosis, and dilated cardiomyopathy (**Table 1**, **Supplementary Table 2**).

**Table 1.** Renal and ureteral phenotypes of the CAKUT cohort. RHD: renal hypo/dysplasia; VUR, vesicoureteral reflux. Other urinary abnormalities include hyperuricemia, reflux nephropathy, posterior urethral valve, hypospadias, ureteropelvic junction obstruction, ureteral stricture, and ureterocyst. Dual renal and urinary phenotypes are counted as separate entries.

| Renal and ureteral phenotypes | Total cases | Sex (male, female) | Bilateral | VUR-associated | With other urinary abnormalities | With extra-renal phenotypes |
| --- | --- | --- | --- | --- | --- | --- |
| Total cohort | 80 | 58, 22 | 61/80 | 22/80 | 13/80 | 35/80 |
| RHD | 45 | 32, 13 | 40/45 | 15/45 | 8/45 | 21/45 |
| Renal cystic dysplasia | 12 | 7, 5 | 8/12 | 3/12 | 1/12 | 6/12 |
| Duplex kidney | 1 | 0, 1 | - | - | 1/1 | - |
| Agenesis | 3 | 3, 0 | 1/3 | 2/3 | - | 1/3 |
| Hydronephrosis | 12 | 11, 1 | 6/12 | 1/12 | 3/12 | 4/12 |
| Renal ectopia or horseshoe kidney | 4 | 2, 2 | 2/4 | 1/4 | - | 2/4 |
| Isolated VUR | 10 | 8, 2 | 7/10 | - | 3/10 | 4/10 |

The median follow-up period was 3.5 years (IQR: 2.1–6.5). During this time, one patient was lost to follow-up and another patient died of heart failure. A total of 56.3% (45/80) patients progressed to chronic kidney disease (CKD) stages II–V, with 23.8% (19/80) progressing to ESKD and necessitated kidney transplantation. At the last follow-up, 23 patients had normal kidney function, while the remaining patients were classified as CKD stage I (n = 10), CKD stage II (n = 15), CKD stage III (n = 15), CKD stage IV (n = 6), or ESKD (n = 9).

Among them, raw data from 78 trio ES samples were available. For the remaining two cases, only the VCF files and phenotypic information of the probands and their parents were accessible.

## Novel candidate genes for CAKUT

To identify additional variants potentially contributing to CAKUT, raw trio ES data from 78 probands, with negative diagnostic genetic testing, were available and re-analyzed (**Figure 1**). Kinship verification led to the exclusion of five samples due to discordant parentage, as assessed by identity-by-decent (IBD) analysis using KING(*54*). Therefore, a total of 73 families were retained for subsequent analyses. Using a joint variant calling strategy(*55*) (**Methods**), we identified a total of 297,836 high-quality variants (272,270 single nucleotide variants (SNVs), and 25,566 small insertions and deletions (indels)), including 185 de novo variants (86 SNVs and 99 indels) and 297,651 inherited variants (272,184 SNVs and 25,467 indels) (**Figure 1**). On average, each proband harbored 2.9 de novo variants (95% confidence interval (CI): 2.5–3.3; **Supplementary Table 3**).

**Figure 1.**
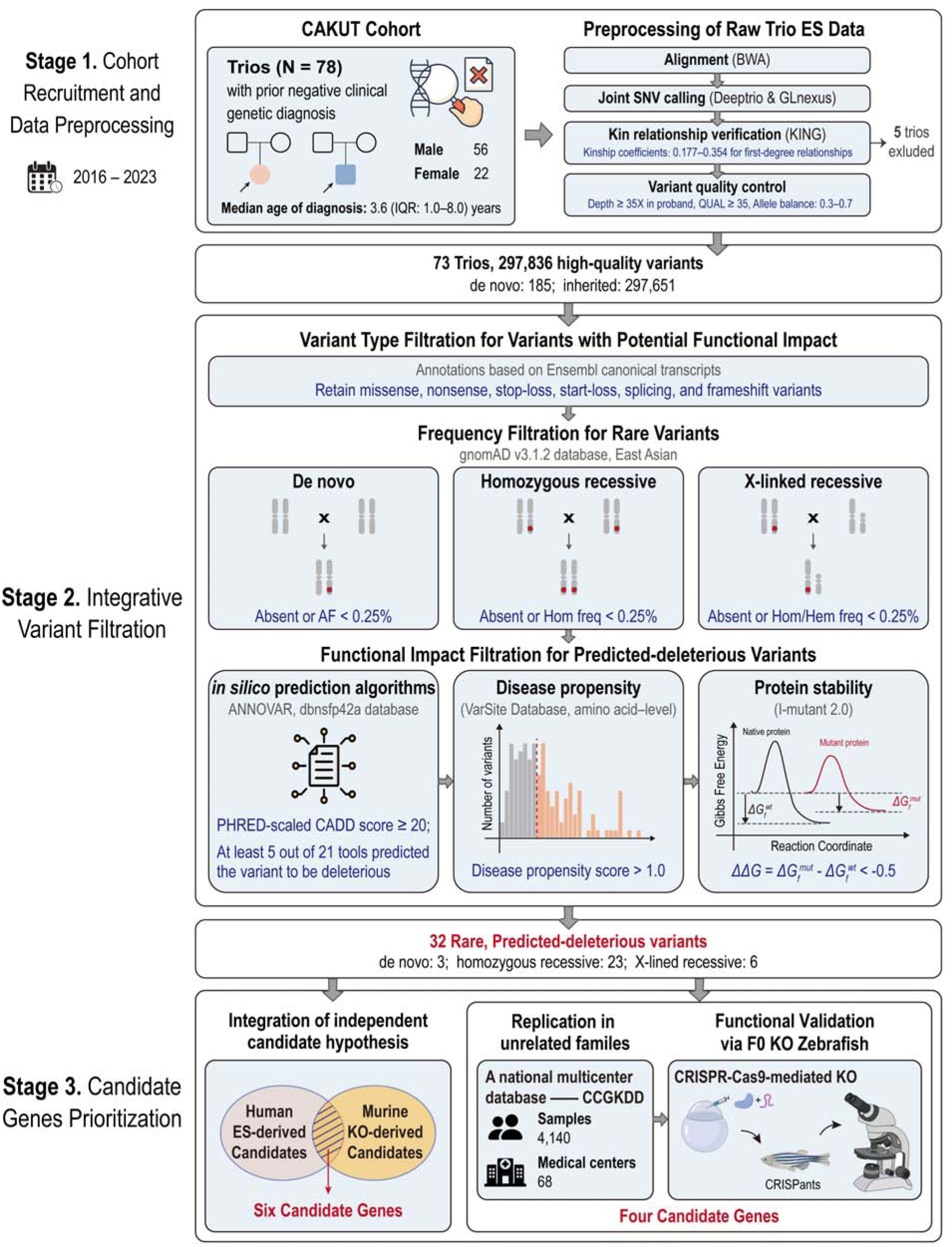
Schematic overview of the study design and integrative analytical framework. In the cohort recruitment and data preprocessing stage, we enrolled 78 trios with congenital anomalies of the kidney and urinary tract (CAKUT) with prior negative clinical genetic diagnoses (median age of diagnosis: 3.6 years; 56 males, 22 females). We processed raw trio exome sequencing (ES) data through alignment and joint variant calling. After kinship verification and variant quality control, five trios were excluded from downstream analyses. Thus, a total of 297,836 high-quality variants were identified across 73 trios, comprising 185 de novo variants and 297,651 inherited variants. In the integrative variant filtration stage, variants with potential functional impact (missense, nonsense, stop-loss, start-loss, splicing, and frameshift variants) were retained based on matched annotation from NCBI and EBI (MANE) transcript annotations. After that, rare variants were filtered across three modes of inheritance—de novo, homozygous recessive, and X-linked recessive—based on the gnomAD database (version 3.1.2) using East Asian samples. Subsequently, predicted deleterious variants were prioritized by simultaneously meeting predefined thresholds across *in silico* pathogenicity scores, amino acid–level disease propensity, and protein stability (quantified by the predicted free energy change, denoted as ΔΔG). In the candidate genes prioritization stage, genes were prioritized via two distinct strategies. Integration of independent evidence from both murine knockout (KO) models and human ES analyses prioritized six candidate genes. In addition, replication in unrelated families from a multicenter cohort, coupled with functional validation via CRISPR-Cas9-mediated KO in F0 zebrafish, yielded four final candidates. AF, allele frequency; Hom freq, homozygote frequency; Hem freq, hemizygote frequency.

In addition to applying population-level frequency-based filtration, we retain variants predicted to have significant functional consequences based on multiple complementary predictors, including machine learning-based pathogenicity scores, amino acid–level disease propensity, and protein stability change (**Figure 1**, **Methods**). This integrative variant filtration approach yielded 32 candidate variants, comprising three de novo SNVs (1.6% out of 185 high-quality calls), 23 homozygous recessive SNVs (0.02% out of 92,724 high-quality calls), and six X-linked recessive SNVs (0.3% out of 1,785 high-quality calls) (**Figure 1**, **Supplementary Table 4**). On average, approximately 0.04 de novo and 0.3 homozygous recessive rare, predicted deleterious variants were identified per proband.

The identified candidate disease-causing variants were distributed across 32 candidate genes. All these genes showed detectable expression in at least one transcriptomic dataset derived from renal and urinary tract tissues in human and mouse, as determined by both bulk and single-nucleus RNA sequencing datasets(*56–65*) (**Figure 2a, Supplementary Figure 1, Supplementary Table 5**). Immunohistochemistry profiling, sourced from the Human Protein Atlas(*66*), further demonstrated protein expression of 25 candidate genes in kidney and bladder tissues, with distinct staining patterns across cells in glomeruli, tubules, and urothelium (**Figure 2a**). None of the 32 ES-derived candidates has been previously implicated in human CAKUT, based on a curated reference set of 185 human genes(*4*)—comprising 54 monogenic CAKUT genes and 131 genes implicated in syndromic disorders exhibiting facultative CAKUT manifestations—and the Online Mendelian Inheritance in Man (OMIM) database(*67*) (as of Nov 22, 2024).

**Figure 2.**
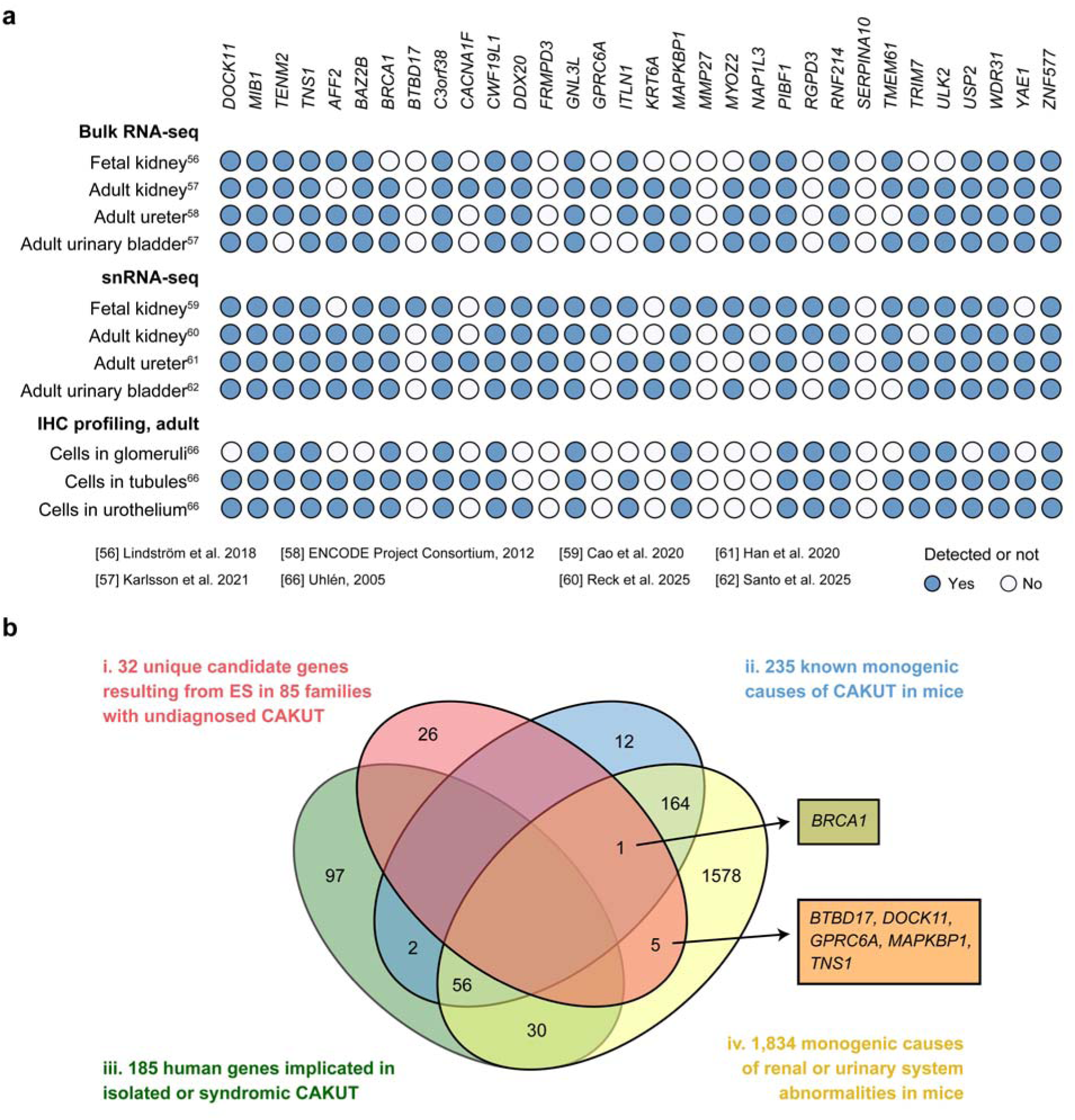
Expression profiles and murine-model-based prioritization of the 32 ES-derived candidate genes. **a.** Expression profiles of 32 exome sequencing (ES)-derived candidate genes across human fetal and adult kidney, urinary bladder, and ureter tissues. Data were sourced from bulk RNA-seq, single-nucleus RNA-seq (snRNA-seq), and immunohistochemistry (IHC) datasets(*56–62*). Each column represents a gene candidate and each row corresponds to an expression profile. Filled circles indicate detected expression at transcriptomic or proteomic level, while open circles indicate absence of detection. **b.** Venn diagram highlights six of the 32 candidate genes for human congenital anomalies of the kidney and urinary tract (CAKUT), identified through integration with murine model-derived candidate genes linked to kidney abnormalities. Venn diagram illustrating intersections among four gene sets: (i, red) 32 unique candidate genes identified by ES in 73 trios with prior negative clinical genetic testing; (ii, blue) 235 known monogenic causes of CAKUT in murine models; (iii, green) 185 human genes implicated in isolated or syndromic CAKUT; and (iv, yellow) 1,834 murine genes whose knockout results in renal or urinary system abnormalities. Of the 185 human genes implicated in isolated or syndromic CAKUT (iii, green), 58 (31%) overlap with the murine CAKUT genes (ii, blue) and 86 (46%) overlap with the broader murine renal/urinary abnormality set (iv, yellow). Among the 32 ES-derived candidate genes (i, red), one (3%) overlaps with the murine CAKUT set (ii, blue) and five (16%) overlap with the broader murine renal/urinary abnormality set (iv, yellow). Six representative overlapping genes are indicated in boxes.

Independent orthogonal layers of evidence are critical to prioritize potential CAKUT candidate genes. Building on previously established framework to prioritize novel candidate genes for kidney diseases(*68*, *69*), we evaluated ES-derived candidate genes (n = 32, **Figure 2b, red**) for roles in kidney development and function by integrating evidences from two resources: a recent systematic review(*4*) of monogenic causes of murine CAKUT (n = 235, **Figure 2b, blue**), representing candidate genes for human CAKUT, and the candidate genes derived from the Mouse Genome Informatics database(*70*) (version 6.24) which catalogs all genes whose knockout produces renal or urinary system abnormalities (n = 1,834, **Figure 2b, yellow**). Six out of 32 ES-derived candidate genes including *BRCA1*, *BTBD17*, *DOCK11*, *GPRC6A*, *MAPKBP1*, and *TNS1* were implicated in renal development and function through evidence from knockout mouse models. For example, *Brca1* has been identified as a monogenic cause of murine CAKUT, with targeted knockout mice displaying ureteral obstruction that mirrors hallmark features of human CAKUT. Similarly, knockout of *Btbd17, Dock11, Gprc6a, Mapkbp1,* and *Tns1* in mice resulted in renal or urinary system abnormalities, including kidney cysts (*Tns1*), abnormal kidney morphology (*Tns1*), increased kidney weight (*Dock11*, *Mapkbp1*), and elevated urinary calcium levels (*Gprc6a*), supporting their potential relevance to human CAKUT.

To further elucidate functional pathways associated with these 32 candidates in our study, they were integrated with 185 known human genes implicated in isolated or syndromic CAKUT (**Figure 2b, green**) to ensure a statistically significant pathway enrichment. Gene ontology (GO) enrichment analysis was performed using Metascape(*71*) (**Supplementary Table 6**). Overall, 23 ES-derived candidate genes were potentially linked to renal or urinary morphology. These candidates were significantly enriched in 193 distinct GO terms (**Benjamini-Hochberg-adjusted *P* value <0.1, Hypergeometric test, Figure 3a**), together with 11 previously reported CAKUT-associated human genes, which are established genetic markers for renal or urinary system morphogenesis (catalogued by a prior review(*72*), **Supplementary Table 7**), indicating a shared involvement of these genes in core biological pathways underlying renal and urinary tract development. Some of these pathways have been implicated in CAKUT pathogenesis, such as Notch signaling(*27*) and epithelial or tube morphogenesis(*22*, *73*) (*MIB1*), MAPK cascade signaling(*31*) (*CACNA1F*, *MAPKBP1*), cilium assembly(*8*) (*MIB1*, *PIBF1*), RAC1 GTPase cycle (*DOCK11*), as well as PI3K-Akt signaling(*74*) (*BRCA1*) (**Figure 3b**).

**Figure 3.**
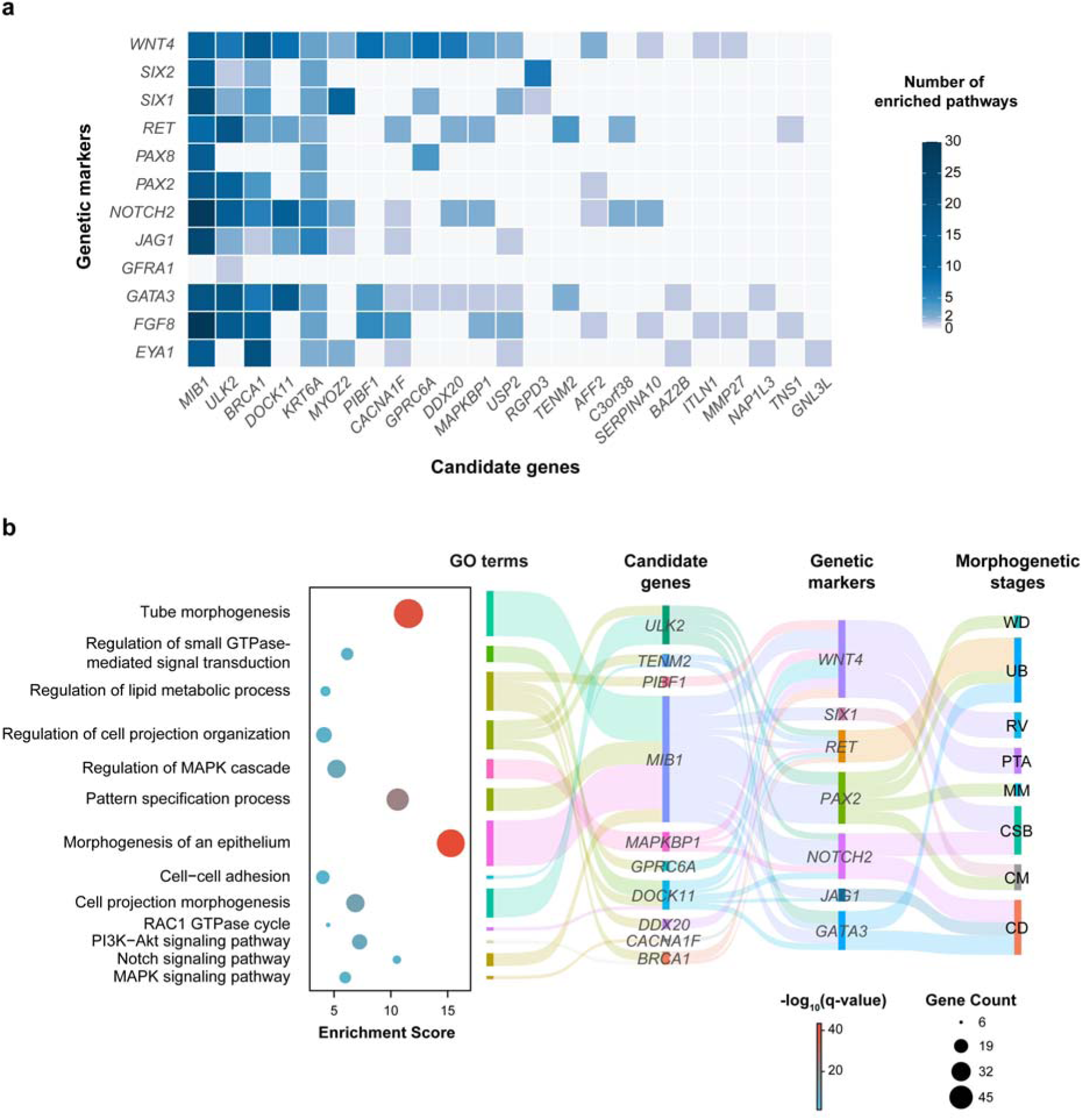
GO enrichment analysis of 32 ES-derived candidate genes integrated with 185 known CAKUT genes. **a.** Heatmap illustrates the co-occurrence matrix of enriched pathways shared by 23 of the 32 exome sequencing (ES)-derived candidate genes and 11 of the genetic markers involved in renal or urinary morphogenetic stages (as reviewed by Khoshdel Rad N et al.(*72*)). Each column represents a gene candidate and each row corresponds to a genetic marker. Colors correspond to the number of enriched pathways, with darker shades indicating a greater number of involving pathways. **b**. The bubble plot illustrates enriched pathways potentially contributing to congenital anomalies of the kidney and urinary tract (CAKUT) pathogenesis. The X-axis represents the enrichment score, while the Y-axis lists the pathway names. The color and size of each circle indicate Benjamini-Hochberg-adjusted *P* value and the number of genes within each pathway, respectively. The sankey diagram maps ES-derived candidate genes to morphogenetic stages of the ureteric and nephron lineages, based on GO enrichment together with the genetic markers of these stages (catalogued by a previous review(*72*), **Supplementary Table 7**). WD, wolffian duct; UB, ureteric bud; RV, renal vesicle; PTA, pre-tubular aggregate; MM, metanephric mesenchyme; CSB, comma-shaped body; CM, cap mesenchyme; CD, collecting duct.

Replication of rare, predicted deleterious variants in unrelated families or independent cohorts has been considered strong gene-level evidence(*18*, *75*), consistent with previously published multicenter and replication studies on genetic kidney disease(*76*, *77*). We screened for additional probands carrying variants in these candidate genes, including only those whose familial inheritance patterns were consistent with the modes observed in our cohort based on the multicenter CCGKDD database(*49*), comprising 1,189 CAKUT patients from 68 medical centers across China. This yielded two additional probands with CAKUT harboring missense variants in the *DOCK11* and *MIB1* gene. Ultimately, six rare, predicted deleterious variants in four candidate genes (*DOCK11*, *MIB1*, *TENM2*, *TNS1*) were identified, each gene recurring in two unrelated probands (**Supplementary Table 8**).

We next delineated the variant spectra and functional context of each gene (**Figure 4a**). The *MIB1* (NM_020774) harbored a de novo nonsense variant, c.2245A>T (p.K749X), and a de novo missense variant, c.1511C>T (p.A504V), neither of which was reported in gnomAD(*78*). The remaining three genes exclusively carried missense variants, each with a hemizygous or homozygous frequency <0.0025 in the gnomAD(*78*) database. In *DOCK11* (NM_144658), two distinct hemizygous variants—c.C2011T (p.R671W) and c.T3391A (p.Y1131N)—were identified in two unrelated male probands. In addition, autosomal-recessive homozygous missense variants were observed in *TENM2* (NM_001395460, c.G5032A, p.G1678S) and *TNS1* (NM_001387777, c.C2950T, p.R984W), each recurring in two unrelated probands.

**Figure 4.**
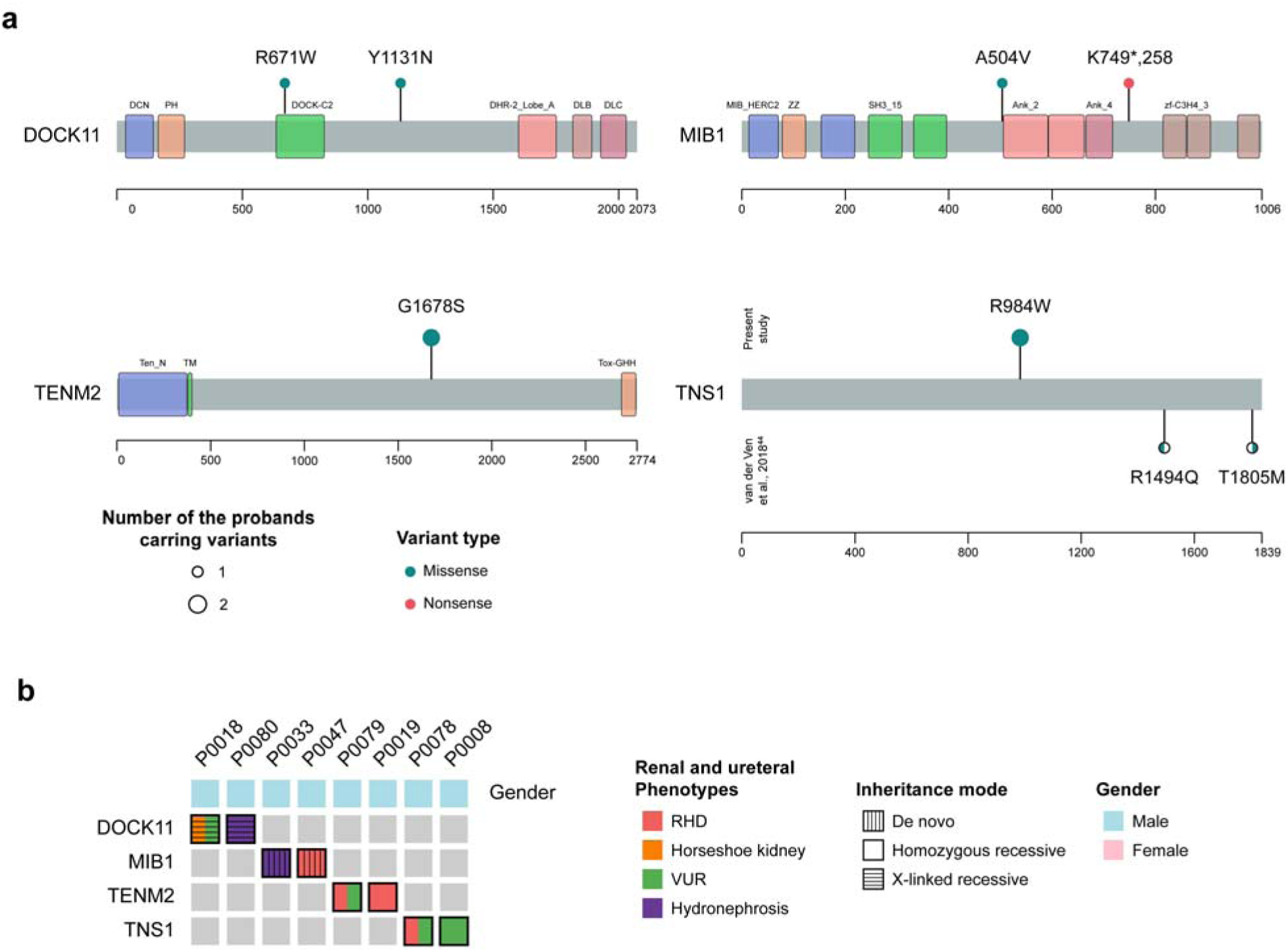
Overview of rare, predicted deleterious variants in four CAKUT candidate genes and the phenotype spectra of corresponding probands. **a.** Distribution of the variants in the protein domain context annotated by InterPro database(*137*). Each panel shows one candidate gene, with its protein domains annotated. Samples from our cohort are displayed above each track. Shown below the *TNS1* track, a compound heterozygous candidate deleterious variant—comprising c.G4481A (p.R1494Q) and c.C5414T (p.T1805M)—was identified in a single congenital anomalies of the kidney and urinary tract (CAKUT) case, as submitted to the Human Gene Mutation Database (accession number: CM1812836) by van der Van et al.(*43*). Lollipop markers represent the positions and types of variants, color-coded by variant type, with circle size denoting the number of probands harboring specific variants. The compound heterozygous variant in *TNS1* is depicted as half-circles. **b.** Phenotypic spectrum of renal and ureteral abnormalities associated with four candidate genes. Each column represents a proband, while each row corresponds to a gene candidate. Colors represent the renal and ureteral phenotypes or the gender. RHD, renal hypo/dysplasia; VUR, vesicoureteral reflux.

All deleterious variants exhibited a high disease propensity score (>1, by VarSite(*79*)) or were predicted to reduce protein stability (ΔΔG <−0.5) using I-Mutant2.0(*80*), with several located within essential functional domains of the respective genes (**Figure 4a, Supplementary Table 8**). For example, the *MIB1* p.K749X truncation removes the C-terminal 258 amino acids, containing three C3HC4-type zinc-finger domains (amino acids 815–1002) essential for its E3 ligase activity(*81*), while p.A504V perturbs a helix-capping alanine, predicted to impair α-helix nucleation in the ankyrin repeats domains (amino acids 506–663) according to previous studies on amino acid secondary structure propensities(*82–85*). In *DOCK11*, p.R671W was located in the C2 domain, which mediates membrane-lipid binding and anchors the protein to microtubule-and actin-based cytoskeletal scaffolds(*86*).

Given the pronounced genetic and clinical heterogeneity observed in CAKUT(*4*, *39–41*), we next characterized the phenotypic spectra associated with each of the four candidate genes (**Figure 4b**). Among *MIB1* variant carriers, the individual (P0033) harboring the de novo nonsense variant (p.K749X) presented with congenital hydronephrosis, whereas the carrier of the missense variant (p.A504V) showed bilateral renal dysplasia. For *DOCK11*, the p.Y1131N carrier (P0018) exhibited hydronephrosis, while the p.R671W carrier had a horseshoe kidney accompanied by bilateral VUR. In the *TENM2* and *TNS1* gene, pairs of unrelated probands shared identical variants. Both probands carrying the c.G5032A (p.G1678S) variant in *TENM2* exhibited renal hypo/dysplasia: left renal dysplasia with bilateral VUR (P0079), and bilateral small kidneys (P0019). In addition, for *TNS1*, two unrelated probands carried the identical homozygous missense variant (c.C2575T, p.R859W) and both presented with bilateral VUR.

We employed a zebrafish model to investigate the potential functional roles of these four candidate genes in renal development and function. Although *TNS1* has been proposed as a candidate gene for CAKUT based on the identification of a compound heterozygous variant—c.G4481A (p.R1494Q) and c.C5414T (p.T1805M)—in a single proband in a prior study(*43*) (**Figure 4a**), its functional role in CAKUT pathogenesis has not yet been characterized.

### CRISPR-Cas9-mediated functional validation of novel candidates in F0 zebrafish

To elucidate the functional roles of the four strong candidate genes implicated in CAKUT pathogenesis, CRISPR-Cas9 gene editing technology was employed to generate zebrafish knockout models and subsequently characterized the knockout embryo phenotypes (**Figure 5a**). Given the kidney’s critical role in regulating water and electrolyte homeostasis, previous studies identified a strong association between renal dysfunction and pericardial edema(*87–89*), leading to the use of the pericardial edema phenotype as a screening tool for identifying candidate genes involved in human kidney diseases.

**Figure 5.**
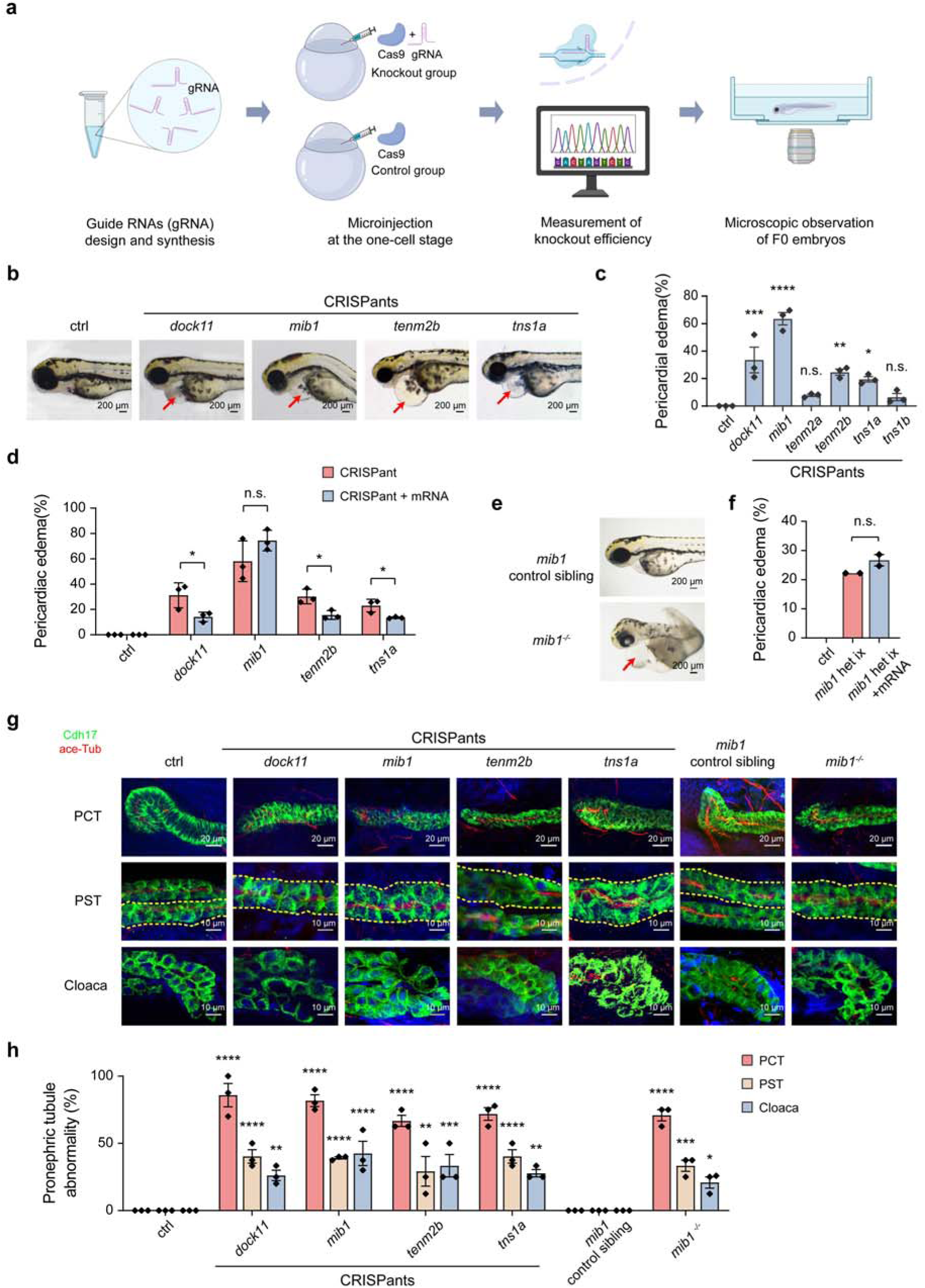
Rapid screening of CAKUT gene candidates using F0 knockout zebrafish (referred to as CRISPants). **a.** Experimental design for zebrafish F0 CRISPants construction. **b.** Representative images of pericardial edema in zebrafish F0 CRISPants, which was injected with Cas9 protein and 4gRNA mixtures, with no edema observed in the control. Red arrows indicate the locations of pericardial edema. **c.** Pericardial edema rates in zebrafish CRISPants, with three independent biological replicates performed per gene. In each replicate, more than 50 zebrafish embryos were observed. **d.** Pericardial edema rates in zebrafish CRISPants (red bars) and the rescued groups which were co-injected with respective wild-type mRNA (blue bars). **e.** Representative images of pericardial edema in *mib1^-/-^* mutants, with no edema observed in *mib1* control siblings. The red arrow indicates the location of pericardial edema. **f.** Pericardial edema rates in embryos from *mib1* heterozygote in-cross with or without wild-type mRNA overexpression. **g.** Representative images of renal tubule segment immunostaining in control and zebrafish CRISPants, with Cadherin-17 (Cdh17, green) and acetylated-Tubulin (ace-Tub, red) indicated. In the anterior region, the proximal convoluted tubules (PCT) of CRISPants exhibit an absence of the typical convoluted structure. In the middle region, the proximal straight tubules (PST) display marked dilation, while the posterior segment near the cloaca shows a loss of the normal bending architecture. **h.** Percentages of abnormal pronephric tubules, including abnormalities in PCT, PST or cloaca, were assessed with three independent biological replicates per gene, and each containing more than eight embryos. Representative images in **b** and **e** are shown in lateral view, with anterior to the left. All embryos were examined and imaged at 3 days post-fertilization. All data are represented as the mean ± SEM. *P* values were calculated by one-way ANOVA for **c**, two-way ANOVA for **d** and **h**, and by two-tailed t-test for **f**. \**P* <0.05; \*\**P* <0.01; \*\*\**P* <0.001; \*\*\*\**P* <0.0001; n.s., non-significant.

A total of six zebrafish genes, including *dock11, mib1, tenm2a, tenm2b, tns1a,* and *tns1b*—orthologous to four human candidate genes—were disrupted by one-cell-stage injection of Cas9 proteins and a mixture of four guide RNAs (gRNAs) targeting each gene. Significant increases (***P* value <0.05, one-way analysis of variance (ANOVA)**) in the percentage of pericardial edema were noted in *dock11, mib1, tenm2b,* or *tns1a* knockout, suggesting renal impairment in these embryos (**Figure 5b–c**). To verify the specificity of the pericardial edema phenotypes in F0 knockout embryos (hereafter referred to as CRISPants), rescue experiments were carried out by co-injection of wild-type mRNA into the respective CRISPants. The pericardial edema phenotypes were significantly rescued with mRNA overexpression in *dock11*, *tenm2b* or *tns1a* CRISPants (***P* value <0.05, two-way ANOVA**), but not in *mib1* CRISPants (**Figure 5d**). To further validate the phenotype specificity of *mib1* CRISPants, we generated a second batch using independent Cas9/gRNA pairs targeting non-overlapping sites and again observed significant pericardial edema phenotypes (**Supplementary Figure. 2a–d**). Furthermore, *mib1^-/-^* mutants also exhibited pericardial edema phenotypes, which also could not be rescued by overexpression of wild-type *mib1* mRNA (**Figure 5e–f**). These observations suggest that *mib1* is required to prevent pericardial edema formation. To summarize, knockouts of *dock11, mib1, tenm2b,* or *tns1a* result in pericardial edema phenotypes in the zebrafish embryos.

We evaluated kidney morphology in these four CRISPants. By Cadherin-17 immunostaining, which labels the pronephric tubules, we found that all four CRISPants exhibited significant morphological abnormalities in the pronephros (***P* value <0.05, two-way ANOVA**), including loss of convoluted structure in proximal convoluted tubule (PCT), tubular dilation, or abnormal cloaca (**Figure 5g–h**). Similar to *mib1* CRISPants, *mib1^-/-^* mutants also showed defects in PCT and tubular dilation (**Figure 5g–h**). Thus, loss of *dock11, mib1, tenm2b,* or *tns1a* lead to defects in renal development in the zebrafish embryos.

We assessed the glomerular filtration function of the CRISPants by evaluation of the FITC-conjugated dextran (Dextran-FITC) clearance efficiency. Dextran-FITC was injected into the sinus venosus of zebrafish hearts, which would gradually be cleared during renal filtration. In wild-type embryos, Dextran-FITC was almost completely cleared after 24 hpi. In contrast, Dextran-FITC still remained in the pericardial cavity in all four CRISPants as well as the *mib1^-/-^*mutants (**Supplementary Figure. 3a–b**), indicating impaired renal filtration function. Therefore, our results indicate that these four candidate genes play essential roles in the zebrafish renal filtration function.

## Discussion

In this study, we performed a comprehensive investigation of children with CAKUT who had negative findings from routine clinical genetic testing and presented a practical analytical pipeline for the systematic identification and prioritization of candidate variants and genes. Through a retrospectively analysis of trio ES data, we identified four candidate genes, and subsequently validated their roles in renal development and function using CRISPR-Cas9-mediated F0 knockout zebrafish models.

As the leading cause of ESKD in children and adolescents, the genetic etiologies of CAKUT have been extensively studied(*1*, *43*, *44*, *53*, *90–95*), however, most investigations have focused on cohorts of European ancestry. By integrating samples from 68 medical centers across China, we identified four novel candidate genes for CAKUT in a Chinese pediatric cohort, with replication in unrelated families, thereby expanding the known genetic landscape of the disease.

Our GO-based analyses, complemented by a targeted literature survey, suggest that these four candidate genes may influence CAKUT risk via diverse pathways, including both well-characterized processes and mechanisms that remain poorly understood.

Prior work identified MIB1 as an E3 ligase to catalyze the ubiquitylation and endocytosis of the ligands of the Notch signaling(*96*), dysregulation of which can force early exit of nephron progenitors from their niche(*27*). In addition, MIB1 determines proximal fates during the comma-and S-shaped body stages in the developing nephrons in mice(*27*). As a guanine nucleotide exchange factor, deficiency in DOCK11 was reported to disrupt the activation of CDC42(*97*). Given that conditional deletion of *Cdc42* in the UB and collecting-duct lineage has been shown to cause severe UB branching defects and abnormalities in epithelial polarity and lumen morphogenesis(*98*), these observations together suggest a potential role for DOCK11 in UB branching. Previous studies have implicated the *TNS1* gene in Mek/Erk signaling(*99*). In murine knockout models, *Tns1* deficiency hyperactivated Mek/Erk signaling and disrupted the integrity of cell–cell junction in lumenogenesis, which underpins UB branching morphogenesis(*99*). In contrast, the contribution of TENM2 in CAKUT still needs to be characterized. TENM2 plays a pivotal role in neurodevelopment by regulating neuronal connectivity(*100*); specifically, it mediates axonal growth cone steering(*101*) and induces the formation of either inhibitory or excitatory synapses(*102*). To date, no genetic disorder has been directly associated with TENM2 according to the OMIM (as of Dec 31st, 2025) or Human Phenotype Ontology (HPO; version 2.1.2) database. Nevertheless, TENM2 represents a compelling candidate for CAKUT pathogenesis, as its roles in cell–cell adhesion (GO:0098609) could plausibly affect UB morphogenesis, including branching patterning and epithelial cell rearrangements in the caudal Wolffian duct—an epithelial tube derived from intermediate mesoderm from which the tip of the primary ureteric bud emerges(*103*, *104*). Perturbations in these processes can lead to CAKUT phenotypes, such as RHD or congenital obstructive uropathy(*16*, *103*). Together, these observations support the four candidate genes as high-confidence contributors to CAKUT, although further studies are required to elucidate their etiological role.

Consistent with previous studies(*4*, *39–41*), we observed the complex genetic landscape of CAKUT, revealing significant phenotypic heterogeneity even among patients harboring variants in the same genes (**Figure 4**). For example, we observed that probands with variants in the *MIB1* gene had distinct renal structural abnormalities—hydronephrosis versus bilateral renal dysplasia. A parallel spectrum of phenotypic variability was seen for *DOCK11*. This phenotypic discrepancy can be explained by several factors, including the exact nature and location of the variants within a gene(*105–107*) or existence of genetic modifiers(*105*, *108–110*), as well as environmental factors such as infections(*111*), diet(*112*), and drug exposure(*113*).

Functional validation of novel candidate genes was performed using CRISPR-Cas9-mediated in F0 zebrafish for the corresponding orthologs of each candidate gene since zebrafish have been established as a model for kidney function and disease(*114*). Increases in pericardial edema rates, abnormal renal tubular morphology and impairment in renal filtration function, were noted in CRISPants of these four genes. In addition, we co-injected the wild-type mRNA in corresponding CRISPants to determine whether restoring their expression could rescue the increased pericardial edema rates observed in the mutant embryos. While co-injection of *dock11*, *tenm2b*, and *tns1a* successfully rescued the abnormalities in the CRISPants, mRNA injections failed to rescue the elevated pericardial edema rates observed in CRISPants as well as mutants of *mib1*, a component of the Notch signaling pathway(*115*). In renal epithelial cells, Notch signaling is distributed in a “salt-and-pepper” pattern due to lateral inhibition(*116*). When *mib1* expression was restored, it could lead to a global activation of Notch signaling, yet not restoring this necessary mosaic pattern and ultimately failing to restore renal function in the *mib1* knockouts.

This study has certain limitations. First, it focused on coding-region variants, without analyzing CNVs, tandem repeats, or non-coding variants, limiting the scope of genetic insights. Secondly, although supporting evidence for candidate genes based on unrelated probands was obtained from multicenter databases, further validation in fully independent cohorts is still required to reduce the risk of overinterpretation and to strengthen genotype–phenotype associations. Moreover, zebrafish models revealed discrepancies, potentially due to structural and functional differences between their pronephros and mammalian metanephros, though zebrafish and human kidneys share conserved segmental patterns. Additionally, based on observations of pronephros tubule morphogenesis in zebrafish models, we mainly validate candidate genes essential for kidney morphogenesis, which led to less thorough examination of functionally impactful genes. Future studies integrating long-read sequencing, mammalian models, and human kidney organoids could address these gaps and provide a more comprehensive understanding of genetic contributions to human CAKUT.

In summary, we systematically reanalyzed trio ES data from CAKUT probands with prior negative clinical genetic testing, with replication evidence in unrelated families from CCGKDD, a national multicenter database across China. This approach enabled identification of novel candidate genes in CAKUT pathogenesis, thereby expanding the genetic landscape of pediatric kidney and urinary tract anomalies.

## Materials and Methods

### Patient cohort and clinical phenotypes

We enrolled 80 probands (58 males, 22 females) diagnosed with CAKUT based on kidney imaging studies. All biological parents of the probands were screened and excluded from a CAKUT diagnosis based on renal imaging studies and urinalysis. Data on demographics, genetic diagnosis, genetic test methods, CAKUT and extrarenal phenotypes were collected for further analysis.

The primary diagnosis of kidney disease was established by each patient’s pediatric nephrologist or urologist based on clinical evaluation and imaging findings. CAKUT was defined as any abnormality in the number, size, shape, or anatomical position of the kidneys or urinary tract, including renal agenesis, RHD, multicystic dysplastic kidney, hydronephrosis, VUR, ectopic or horseshoe kidney, duplex collecting system, posterior urethral valves, ureteropelvic junction obstruction, hydroureter, and ureterovesical junction obstruction.

Glomerular filtration rate was classified as G1 (≥90 mL/min/1.73 m²), G2 (60–89), G3 (30–59), G4 (15–29), and G5 (<15). Quantitative variables were summarized as the median and interquartile range (IQR).

All patients were sourced from a national multicenter registration network (CCGKDD, www.ccgkdd.com.cn), which was officially launched in 2017. Led by local experts from the National Pediatric Medical Center of China, the database integrates clinical information and research results from 68 centers (**Supplementary Note**) in China, whose capabilities in genetic diagnosis technology are qualified, allowing for early and accurate diagnosis and treatment. Clinical features of the included renal diseases included five major categories: (a) CAKUT; (b) glomerulopathy (including steroid-resistant nephrotic syndrome, nephritis, and non-nephrotic proteinuria, and atypical hemolytic uremic syndrome); (c) renal cystic disease (including nephronophthisis, and polycystic kidney disease); (d) renal tubular disease, nephrolithiasis and renal calcinosis; and (e) CKD 3 to 5 stage with unknown origin. As of December 19, 2025, 4,140 samples were registered. Among these, 1,189 samples were diagnosed with CAKUT.

### Clinical Genetic Testing

All probands underwent clinical genetic testing between 2016 and 2023 before the age of 18 years.

Peripheral blood treated with EDTA was collected. Genomic DNA was extracted using the Blood Genome Column Medium Extraction Kit (CWBIO, China) according to the manufacturer’s protocol. DNA quality was assessed using a Qubit 2.0 fluorometer and 0.8% agarose gel electrophoresis before downstream analyses.

Protein-coding exome enrichment was performed using the xGen Exome Research Panel v2.0 (Integrated DNA Technologies, Iowa, USA), comprising 429,826 individually synthesized and quality-controlled probes. This panel targets 39 Mb protein-coding region spanning 19,396 genes within the human genome, covering 51 Mb of end-to-end tiled probe space. High-throughput sequencing was conducted on MGISEQ-T7 sequencer (PE150), operated by Beijing Chigene Translational Medicine Research Center Co., Ltd, 100875, ensuring the sequencing of not less than 99% of the target sequence.

ES-based clinical genetic testing was performed for singletons or trios. The bioinformatics pipeline involved alignment to the GRCh37 reference genome and variant calling for SNVs, indels, and CNVs. Briefly, base quality score recalibration and SNV and indel calling were performed using the Genome Analysis Toolkit (GATK) best practice pipeline(*117*), while CNV analysis was performed using ExomeCNV with default parameters(*118*), which primarily relies on coverage depth information. High-quality variants were filtered based on sequencing depth and quality metrics and were annotated through multiple databases, including dbSNP, ExAC, ESP, HGMD, and Clinvar. The versions of these datasets have changed over time as the samples were sequenced and analyzed between 2016 and 2023. Variant pathogenicity was evaluated according to the standards and guidelines of the American College of Medical Genetics and Genomics.

All samples yielded negative findings with no pathogenic or likely pathogenic variants identified.

### Re-analysis of ES data

Raw data were processed by Trimmomatic version 0.39(*119*) for adapter removal. The paired-end reads were mapped to GRCh38.p14 using Burrows-Wheeler Aligner(*120*) (version 0.7.15-r1140) and the mapping reads were sorted and indexed using SAMtools (version 1.13)(*121*). Polymerase chain reaction duplicates were marked up using picard-tools-2.26.11 through the MarkDuplicates wrapper. The depth module in SAMtools(*121*) was used to calculate sequencing depth on exome targets, while mosdepth (version 0.3.3)(*122*) was employed to determine the average coverage using the ‘--thresholds’ parameter. Each proband reached an average sequencing depth of 42.0X (95% CI: 39.2–44.8), while parental samples averaged 37.4X (95% CI: 35.9–38.9) (**Supplementary Table 9**).

SNVs and indels were identified using DeepTrio v1.4(*55*). Individual variant calls from DeepTrio were stored in gVCF format. Following that, the gVCFs of each family were combined with GLnexus v1.4.1(*55*). Variant-level quality control was performed independently for each trio. Variants were excluded if any family member had missing genotype calls or if they were multi-allelic. Only variants with sufficient sequencing depth (≥35X in the proband, and ≥20X in both parents) and high phred-scaled quality score (VCF QUAL ≥35) were retained. For heterozygous genotypes, allele balance thresholds of 0.3–0.7 were applied(*123*). Additionally, variants that failed to be genotyped (indicated by a period ‘.’) or were multi-allelic were excluded. Variants passing all filters were considered high quality and used in downstream analyses.

Samples were excluded if a discordance in sex, that cannot be explained by aneuploidy, was observed between the genetically inferred sex and the self-reported sex. Genetic sex was inferred using the heterozygosity rate within the X chromosome non-pseudoautosomal region, applying the thresholds of <0.2 for males and >0.2 for females based on the quality control standards in the gnomAD project(*78*). Firstly, high-quality variants were pruned for linkage disequilibrium (LD) using PLINK(*124*) (version 1.9, www.cog-genomics.org/plink/1.9/) with the ‘--indep-pairwise 200 5 0.2’ parameter(*125*). After that, kinship coefficients for each trio were evaluated through IBD analysis using KING (version 2.3.2) (*54*) based on the LD-pruned variant set and the ‘--related --degree 2’ parameters. Default thresholds were applied for kinship coefficients verification: >0.354 for duplicate samples or monozygotic twins, 0.177–0.354 for first-degree relationships, 0.0884–0.177 for second-degree relationships, and 0.0442–0.0884 for third-degree relationships. Five out of 78 trios, exhibiting unexpected proband-parent relationships, were excluded from subsequent analyses.

Variant annotations were performed via ANNOVAR(*126*), using databases including refGene (release 2019Oct24), avsnp150 (release 2017Sep29), gnomad312_genome (release 2022Dec28), dbnsfp42a (release 2021Jul10), and clinvar_20221231 (release 2023Jan05).

To filter out variants with significant functional consequences, we applied integrative filtration strategies. Firstly, we focused on variants with potential functional impact, including missense, nonsense, stop-loss, start-loss, splicing, and frameshift variants, for further investigation. In cases with multiple transcripts, the most deleterious effect was included. Population allele frequencies were obtained from gnomAD82 (version 3.1.2) using East Asian samples. For de novo alterations, rare variants were defined by an allele frequency <0.0025. For homozygous recessive variants, variants with a homozygote frequency >0.0025 were excluded from the candidate set. The retained X-linked recessive variants were subject to a combined frequency of hemizygous and homozygous individuals of <0.0025. Annotations on *in silico* pathogenicity prediction scores were based on dbnsfp42a database (release 2021Jul10). With reference to established filtration strategies from previous studies(*43*, *46*, *127*), a variant was defined as “deleterious” only when they met both of the following criteria: (1) predicted to have deleterious effects by at least five *in silico* prediction algorithms based on ANNOVAR annotation, and (2) possessed a PHRED-scaled CADD(*128*) score ≥20 (GRCh38-v1.6).

After filtering variants based on variant type, population allele frequency, and *in silico* predictions of deleteriousness, we further prioritized candidates by integrating amino acid–level disease-propensity scores from the Varsite database(*79*) and predicted free energy changes (ΔΔG) using I-mutant2.0(*80*). Prediction of protein stability changes upon single amino acid substitutions were calculated based on the translated protein sequences of matched annotation from NCBI and EBI (MANE) transcripts (**Supplementary Table 10**) retrieved via BioMart(*129*).

A positive ΔΔG denotes increased stability; negative values indicate destabilization. Variants were retained in the final set when the disease-propensity score >1.0 and ΔΔG <-0.5.

All filtered variants underwent manual verification to eliminate potential false positives identified by the SNV calling software, according to the alignment generated by the Integrative Genomics Viewer(*130*).

### Replication of gene candidate findings in CCGKDD

Gene candidates were systematically interrogated within the CCGKDD database, which comprises 4,140 individuals from 68 Chinese medical centers, to screen for additional probands carrying either identical or distinct rare, predicted deleterious variants within the same genes. To ensure high-confidence variant calls and maintain data quality across the multi-center dataset, variants with an allelic balance (the ratio of reads aligned at a variant locus that support the alternate allele) lower than 0.3(*123*) were excluded for samples where raw sequencing data were unavailable.

### Analysis of publicly available gene and protein expression data

Following the classification criteria of the Human Protein Atlas(*57*) (HPA, https://www.proteinatlas.org/), we categorize genes as “not detected” with a baseline cutoff of transcripts per million (TPM) <1, with specific adjustments due to distinct normalizing strategies. For adult kidney and urinary bladder tissue transcriptomes sourced from HPA, genes with an nTPM value <1.0 across all samples were classified as “not detected”. In the bulk transcriptome dataset of human developing kidney tissues(*56*), genes were classified as “not detected” if TPM values at all analyzed developmental stages (weeks 9, 11, 13, 17, and 21) were <1.0. For the ENCODE adult ureter dataset, a gene was classified as ’not detected’ only when the upper bound of its TPM confidence interval (TPM_ci_upper_bound) fell below 1(*58*).

Immunohistochemistry profiling data were obtained from the HPA database. Antibody staining levels in the annotated cell types (specifically cells in glomeruli, tubules, and urothelium) were reported as not detected, low, medium, or high, based on conventional immunohistochemistry profiling in selected tissues. This score is based on the combination of the staining intensity and fraction of stained cells(*66*).

The normalized RNA-seq data across developmental stages of the mouse metanephros were sourced from the EGA ArrayExpress database with the accession codes: E-MTAB-6798 (https://www.ebi.ac.uk/arrayexpress/)(63). The gene expression levels were categorized as below Cutoff, low, medium or high based on TPM levels.

Processed single-cell RNA sequencing data of both human and mouse were sourced from CZ CELLxGENE Discover(*131*), a data platform that provides curated and interoperable single-cell datasets, and a gene was classified as “detectable” if it was detected in at least three cells.

### Functional validation studies in zebrafish

Wide-type zebrafish of Tubingen background were maintained according to standard protocol(*132*). Embryos were collected from natural mating and kept at 28.5 °C in E3 solution.

### Mutagenesis by the CRISPR-Cas9 system

Generation of zebrafish CRISPants using the CRISPR-Cas9 system was carried out as previously described(*133*). gRNA targeting sequences (**Supplementary Table 11**) were used for target genes according to the gRNA tables(*133*, *134*) or CRISPRscan website https://www.crisprscan.org/. Both Cas9 protein (NEB cat# M0646M) and four gRNAs were injected into zebrafish embryos at one- to two-cell stages. The mutagenesis efficiency was estimated by Sanger sequencing. The live embryos were photographed with a Nikon SMZ1500 stereomicroscope.

### mRNA injection

Full-length cDNA of *mib1* was amplified from the zebrafish cDNA library, while *dock11*, *tenm2b*, and *tns1a* were synthesized by Beijing Tsingke Biotech Co., Ltd.. All constructs were subcloned into the *pCS2-linker-gfp* vector. The *dock11-gfp*, *mib1-gfp*, *tenm2b-gfp* and *tns1a-gfp* mRNA were transcribed *in vitro* using the mMESSAGE mMACHINE SP6 kit (Ambion) using linearized pCS2-*dock11*-linker-gfp, pCS2-*mib1*-linker-gfp, pCS2-*tenm2b*-linker-gfp, and pCS2-*tns1a-*linker-gfp constructs as templates. Primer sequences used for polymerase chain reaction amplification are provided in **Supplementary Table 11**.

### Immunostaining

Embryos were fixed with Dent’s fixative at -20°C for ≥2 hours, blocked in a PBST buffer containing 10% serum, then incubated with primary antibodies followed by secondary antibodies conjugated with Alexa Flour 488 or Alexa Flour 594. Images were taken with a Nikon A1R confocal microscope.

Primary antibodies were diluted in PBST blocking buffer at following concentrations: anti-ace-tubulin (1:2000, Sigma #6-11B-1), anti-Cdh17 (1:200, customized antibody generated by B&M). Secondary antibodies (Invitrogen) were used at 1:500 dilutions.

### Glomerular filtration rate measurement

FITC-conjugated dextran (Dextran-FITC, 40kD) was injected into the sinus venosus of zebrafish hearts to check its clearance efficiency and evaluate the glomerular filtration function of zebrafish(*135*). The 3 dpf old embryos were injected with 1 nl of 10mg/ml Dextran-FITC solution and photographed at 5 hours post-injection (hpi) and 24 hpi at same conditions using a Nikon SMZ1500 stereomicroscope (Nikon, Tokyo, Japan).

### Statistical Analyses

Statistical analyses were performed using R version 4.1.0, with a statistical significance threshold defined as *P* value <0.05.

## Data Availability

Our study is compliant with the Guidance of the Ministry of Science and Technology (MOST) of China for the Review and Approval of Human Genetic Resources. The raw sequencing data, including BAM files of ES of 78 trios in this study, have been deposited in the Genome Sequence Archive (GSA)(136) in the National Genomics Data Center, Beijing Institute of Genomics, Chinese Academy of Sciences, under accession number HRA010320 (BioProject: PRJCA035766) that can be accessed at http://bigd.big.ac.cn/gsa-human.
To protect participant confidentiality, the data are available to bona fide researchers within the wider scientific community through a controlled access process. Access can be obtained by completing the application form via Genome Sequence Archive (GSA). For detailed guidance on making the data access request, see GSA-Human_Request_Guide_for_Users [https://ngdc.cncb.ac.cn/gsa-human/document/GSA-Human_Request_Guide_for_Users_us.pdf]. The approximate response time for accession requests is about ten working days.

## Data Sharing Statement

Our study is compliant with the Guidance of the Ministry of Science and Technology (MOST) of China for the Review and Approval of Human Genetic Resources. The raw sequencing data, including BAM files of ES of 78 trios in this study, have been deposited in the Genome Sequence Archive (GSA)(*136*) in the National Genomics Data Center, Beijing Institute of Genomics, Chinese Academy of Sciences, under accession number HRA010320 (BioProject: PRJCA035766) that can be accessed at http://bigd.big.ac.cn/gsa-human.

To protect participant confidentiality, the data are available to bona fide researchers within the wider scientific community through a controlled access process. Access can be obtained by completing the application form via Genome Sequence Archive (GSA). For detailed guidance on making the data access request, see GSA-Human_Request_Guide_for_Users [https://ngdc.cncb.ac.cn/gsa-human/document/GSA-Human_Request_Guide_for_Users_us.pdf]. The approximate response time for accession requests is about ten working days.

## List of abbreviations

CAKUT: congenital anomalies of the kidney and urinary tract
VUR: vesicoureteral reflux
ESKD: end-stage kidney disease
UB: ureteric bud
GDNF: glial-derived neurotrophic factor
PI3K-Akt: phosphoinositide 3-kinase-AKT
MAPK: Mitogen-Activated Protein Kinase
cAMP: cyclic adenosine 3′,5′-monophosphate
ES: exome sequencing
RHD: renal hypo/dysplasia
IQR: interquartile range
CCGKDD: Chinese Children Genetic Kidney Disease Database
CKD: chronic kidney disease
SNV: single nucleotide variant
Indel: short insertions and deletion
CI: confidence interval
LD: linkage disequilibrium
IBD: identity-by-decent
TPM: transcripts per million
gRNA: guide RNA
GO: gene ontology
PCT: proximal convoluted tubule
WD: wolffian duct
UB: ureteric bud
RV: renal vesicle
PTA: pre-tubular aggregate
MM: metanephric mesenchyme
CSB: comma-shaped body
CM: cap mesenchyme
CD: collecting duct

## Acknowledgements

The authors thank Prof. Friedhelm Hildebrandt at the Harvard Medical School for his insightful comments on the manuscript. The authors appreciate the affected individuals and their families for their participation and thank Chigene (Beijing) Translational Medicine Research Center Co., Ltd. for their support of sequencing technology. The contributing members of the CCGKDD are listed in the **Supplementary Note**.

## Contributions

S.F. and H.X. conceived the experiments. S.F., H.X., and Y.C. designed the experiments. S.F., Y.C. and Y.H.Z. wrote and revised the manuscript with the help of other authors. H.S. performed bioinformatics analyses including reads mapping, variant calling, annotation, and filtration. Y.C., W.Z. and M.D. conducted the zebrafish experiments. C.W., Y.H.Z. and Qian Shen collected the samples and contributed to clinical diagnoses. J.M., H.L., Qing Sun, H.J.S., J.W., D.X., Y.Z., and M.L. contributed to data collection and clinical information submission. All authors read and approved the manuscript.

## Ethics approval and consent to participate

This research was performed according to the principles of the Helsinki Declaration and complied with all relevant regulations for working with human subjects in China. The Ethics Committee of the School of Life Sciences, Fudan University, Shanghai, China approved the study. Participants were recruited to a project funded by the Ministry of Science and Technology of the People’s Republic of China (2021YFC2500202). Informed consents were approved by all participants. Our study is compliant with the Guidance of the Ministry of Science and Technology (MOST) of China for the Review and Approval of Human Genetic Resources.

