## Supplementary Figures and Supplementary Notes for "Exome Reanalysis Identifies Novel Candidate Genes Associated with Congenital Anomalies of the Kidney and Urinary Tract in China"

### Supplementary Figure

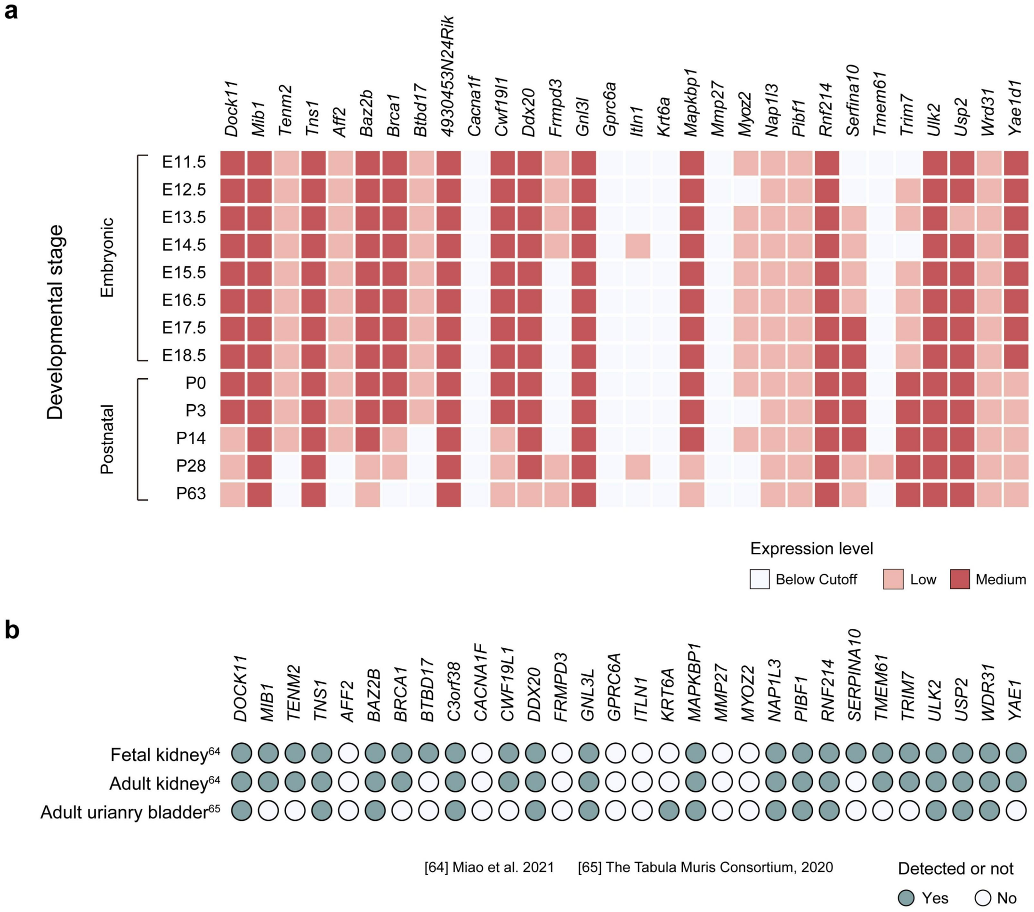


**Supplementary Figure 1. Gene expression profiles of mouse orthologs of ES-derived human candidates. a.** Gene expression profiles of 30 mouse orthologs during metanephros development. The heatmap illustrates the expression patterns of orthologs corresponding to human candidate genes across developmental stages in the mouse metanephros. Each row represents a specific developmental stage, spanning embryonic days (E11.5 – E18.5) to postnatal days (P0 – P63), while each column corresponds to a gene candidate. **b.** Gene expression profiles of 30 mouse orthologs across fetal kidney, adult kidney, and adult urinary bladder tissues. Filled circles indicate confirmed transcriptomic detection, while open circles denote absence of detection. Data for panel **a** were sourced from bulk RNA-seq data(*86*), and data for panel **b** were obtained from single-nucleus RNA-seq data(*87*). The mouse orthologs corresponding to human ES-derived candidate genes were obtained from the Ensembl database, and were listed in **Supplementary Table 8**.

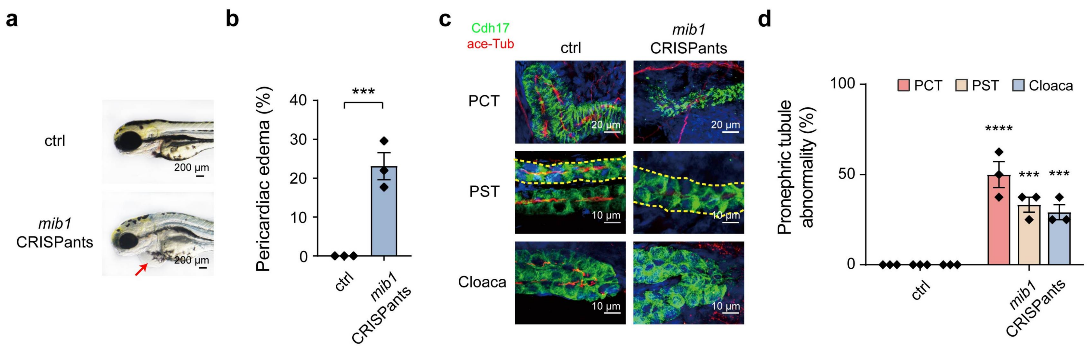


**Supplementary Figure 2. Additional knockout experiments with *mib1*. a.** Representative images of pericardial edema in zebrafish F0 CRISPants, which was injected with Cas9 protein and 4gRNA mixtures, with no edema observed in the control. The red arrow indicates the location of pericardial edema. **b.** Pericardial edema rates in zebrafish CRISPants, with three independent biological replicates performed per gene. In each replicate, more than 50 zebrafish embryos were observed. **c.** Representative images of renal tubule segment immunostaining in control and zebrafish CRISPants, with Cadherin-17 (Cdh17, green) and acetylated-Tubulin (ace-Tub, red) indicated. In the anterior region, the proximal convoluted tubules (PCT) of CRISPants exhibit an absence of the typical convoluted structure. In the middle region, the proximal straight tubules (PST) display marked dilation, while the posterior segment near the cloaca shows a loss of the normal bending architecture. **d.** Percentages of abnormal pronephric tubules, including abnormalities in PCT, PST or cloaca, were assessed with three independent biological replicates per gene, and each containing more than eight embryos. Representative images in **a** are shown in lateral view, with anterior to the left. All embryos were examined and imaged at 3 days post-fertilization. All data are represented as the mean ± SEM. $P$ values were calculated by two-tailed t-test for **b**, two-way ANOVA for **d**. ***$P$ <0.001; ****$P$ <0.0001.

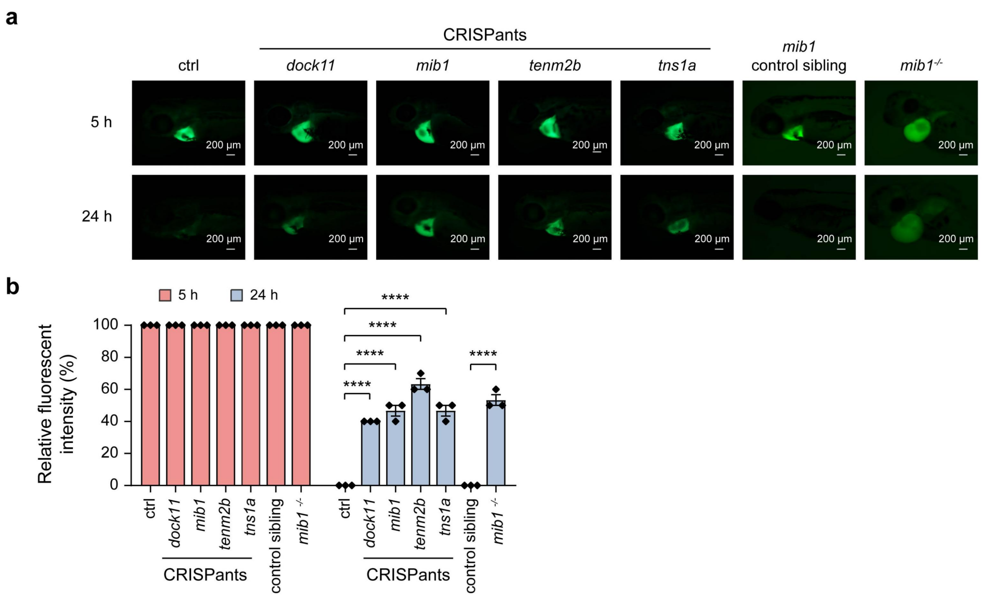


**Supplementary Figure 3. CRISPant embryos exhibit defects in renal filtration function. a.** Representative images of embryos with FITC-conjugated dextran injected into the pericardium at 5 hours post-injection (hpi) and 24 hpi, top and bottom two panels respectively, in control versus zebrafish F0 CRISPants, as well as *mib1* mutants and its control siblings. **b.** Percentages of fluorescent intensity remaining at 5, and 24 hpi in control versus zebrafish F0 CRISPants, as well as *mib1* mutants and its control siblings. Representative images in **a** are shown in lateral view, with anterior to the left. All embryos were examined and imaged at 3 days post-fertilization. All data are represented as the mean ± SEM. *P* values were calculated by one-way ANOVA for **b**. $^{**}P < 0.01$; $^{***}P < 0.001$.

### Supplementary Note

The contributing members of the Chinese Children Genetic Kidney Disease Database (CCGKDD) are listed as follows, and the main persons in charge are indicated.

1. Children's Hospital of Fudan University, Shanghai (Hong Xu, Qian Shen)
2. Beijing Children's Hospital, Capital Medical University (Ying Shen)

3. Shanghai Children's Medical Center, Shanghai Jiao Tong University School of Medicine (Wei Zhou)
4. The Second Hospital of Shandong University (Yulong Wang)
5. First Affiliated Hospital of Henan University of CM (Xia Zhang)
6. Boai Hospital of Zhongshan (Yuling Liu)
7. The First People's Hospital of Yunnan Province (Yan Liu)
8. Anhui Children's Hospital (Fang Deng)
9. Hebei Children's Hospital (Ling Liu)
10. The Affiliated Hospital of Inner Mongolia Medical University (Yanyan Guo)
11. Yantai Yuhuangding Hospital (Weiyan Cai)
12. The First Affiliated Hospital of Anhui Medical University (Xun Xia)
13. Tianjin Children's Hospital (Shaona Song)
14. Chenzhou First People's Hospital (Chenzhou Children's Hospital) (Ling Sun)
15. Henan Children's Hospital (Zhengzhou Children's Hospital) (Cuihua Liu)
16. Children's Hospital Affiliated to Shandong University (Jinan Children's Hospital) (Hongxia Zhang)
17. Qinghai Province Women & Children's Hospital (Jing Cao)
18. The First Affiliated Hospital of Zhengzhou University (Jianjiang Zhang)
19. Xinhua Hospital Affiliated to Shanghai Jiao Tong University School of Medicine (Jing Jin)
20. Chengdu Women's and Children's Central Hospital (Shipin Feng)
21. The University of Hong Kong-Shenzhen Hospital (Rui Liang)
22. Xuzhou Children's Hospital (Huandan Yang)
23. Harbin Children's Hospital (Fang Ning)
24. Urumqi First People's Hospital (Urumqi Children's Hospital) (Lei Yu)
25. North Branch of Ruijin Hospital, Shanghai Jiao Tong University School of Medicine (Yi Yu)
26. Dalian Children's Hospital (Mei Han)
27. Guangzhou First People's Hospital (Zhihong Hao)
28. The first hospital of Jilin University (Qingshan Ma)

29. Lianyungang Maternal and Child Health Hospital (Li Miao)
30. WeiFang Maternal and Child Health Hospital (Jian Gao)
31. Shenzhen Children's Hospital (Xiaojie Gao)
32. The Affiliated Hospital of Qingdao University (Yi Lin)
33. YuYing Children’s Hospital of Wenzhou Medical University (Dexuan Wang)
34. West China Second University Hospital, Sichuan University (Yuhong Tao)
35. Qingdao Women's and Children's Hospital (Qing Sun)
36. Children's Hospital of Chongqing Medical University (Mo Wang)
37. Xi'an Children's Hospital (Ying Bao)
38. Wuxi Children's Hospital (Guoming Li)
39. The Second Hospital of Hebei Medical University (Zanhua Rong)
40. Guiyang Maternal and Child Health Care Hospital (Guiyang Children's Hospital) (Yuhong Li)
41. The Second Xiangya Hospital of Central South University (Xiqiang Dang)
42. Changchun Children's Hospital (Yunkun Han)
43. Peking Union Medical College Hospital (Yanyan He)
44. Xiamen Maternal and Child Health Hospital (Tong Shen)
45. Wuhan Children's Hospital (Xiaowen Wang)
46. Kunming Children's Hospital, Kunming Medical University (Bo Zhao)
47. Shengjing Hospital of China Medical University (Yubin Wu)
48. The First Affiliated Hospital, Sun Yat-sen University (Xiaoyun Jiang)
49. Children's Hospital, Zhejiang University School of Medicine (Jianhua Mao)
50. The First Affiliated Hospital of Xiamen University (Haitao Bai)
51. Shandong Provincial Hospital (Shuzhen Sun)
52. The First Affiliated Hospital of Xinjiang Medical University (Hongtao Zhu)
53. Shanxi Children's Hospital (Lijun Zhao)
54. Xinjiang Uiger Municipal People's Hospital (Feiyan Wang)
55. Children's Hospital of Nanjing Medical University (Aihua Zhang)
56. Maternal and Child Health Hospital of Hubei Province (Xiaolin Wu)
57. General Hospital of Ningxia Medical University (Lijun Liang)

58. The First Affiliated Hospital of Xinxiang Medical University (Ziming Han)
59. Shanghai Children's Hospital, School of Medicine, Shanghai Jiao Tong University (Wenyan Huang)
60. General Hospital of Eastern Theater Command (Nanjing General Hospital of Nanjing Military Region) (Chunlin Gao)
61. Liuzhou People's Hospital, Guangxi (Jiangfeng Peng)
62. Tongji Hospital, Tongji Medical College of HUST (Jianhua Zhou, Liru Qiu)
63. Guangzhou Women and Children's Medical Center (Xia Gao)
64. The First Affiliated Hospital of Jinan University (Guangzhou Overseas Chinese Hospital) (Fang Yang)
65. Foshan Maternal and Child Health Hospital/Foshan Children's Hospital (Yuhua Ma)
66. Children's Hospital of Soochow University (Xiaozhong Li)
67. Xiamen Children's Hospital (Guangbo Li)
68. The 900th Hospital of Joint Logistic Support Force, Fuzhou, China (Xiaojing Nie)
